# The Simple One-step stool processing method for detection of Pulmonary tuberculosis: a study protocol to assess the robustness, stool storage conditions and sampling strategy for global implementation and scale-up

**DOI:** 10.1101/2022.02.04.22270430

**Authors:** Petra de Haas, Bazezew Yenew, Getu Diriba, Misikir Amare, Andrii Slyzkyi, Yohannes Demissie, Bihil Sherefdin, Ahmed Bedru, Endale Mengesha, Zewdu Gashu Dememew, Abebaw Kebede, Muluwork Getahun, Edine Tiemersma, Degu Jerene

## Abstract

**Background:** Xpert MTB/RIF Ultra (Xpert-Ultra) provides timely results with good sensitivity and acceptable specificity with stool samples in children for bacteriological confirmation of tuberculosis (TB). This study aims to optimize the Simple One-Step (SOS) stool processing method for testing stool samples using the Xpert-Ultra in children and adults in selected health facilities in Addis Ababa, Ethiopia. The study is designed to assess the robustness of the SOS stool method, to help fine-tune the practical aspects of performing the test and to provide insights in stool storage conditions and sampling strategies before the method can be implemented and scaled in routine settings in Ethiopia as well as globally.

**Methods and design:** The project “painless optimized diagnosis of TB in Ethiopian children” (PODTEC) will be a cross sectional study where three key experiments will be carried out focusing on 1) sampling strategy to investigate if the Xpert-Ultra *M*.*tuberculosis* (MTB) -positivity rate depends on stool consistency, and if sensitivity can be increased by taking more than one stool sample from the same participant, or doing multiple tests from the same stool sample, 2) storage conditions to determine how long and at what temperature stool can be stored without losing sensitivity, and 3) optimization of sensitivity and robustness of the SOS stool processing method by varying sample processing steps, stool volume, and sampling from the stool-sample reagent mixture.

Stool samples will be collected from participants (children and adults) who are either sputum or naso-gastric aspiration (NGA) and/or stool Xpert-Ultra MTB positive depending on the experiment. Stool samples from these participants, recruited from 22 sites for an ongoing related study, will be utilized for the PODTEC experiments. The sample size is estimated will be 50 participants.

We will use EpiData for data entry and Stata for data analysis purposes. The main analyses will include computing the loss or gain in the Xpert-Ultra MTB positivity rate, and rates of unsuccessful test results. The differences in the positivity rate regarding testing more than one sample per child, different storage, and processing conditions, will be compared to the baseline (on-site) Xpert-Ultra result.

**Ethics and dissemination:** The protocol was reviewed and approved by the Ethical Review Board of the Ethiopian Public Health Institute (EPHI-IRB) (Protocol no EPHI-IRB-234-2020). The study results will be shared with the national TB program and stakeholders to the benefit of further roll out of the test in a routine Ethiopian setting. The results will also be disseminated in peer-reviewed scientific journals.

## Introduction

Approximately 1.09 million children globally fall ill with tuberculosis (TB) each year,, of whom only 399.000 are notified (1). Every day, nearly 700 children die from TB, 80% of them before reaching their fifth birthday. Treatment exists that could prevent nearly all these deaths, but less than 5% of children get treatment as childhood TB is difficult to diagnose (2).

The recent WHO guidelines recommend stool as non-invasive primary diagnostic samples for testing with Xpert and Xpert Ultra for a diagnosis of TB among children (3, 4)based on evidence from multiple studies summarized in three systematic literature reviews ((5), (6), (7). These reviews showed the pooled sensitivities and specificities of stool Xpert against a microbiological (sputum-based) reference standard of between 50 and 67% and 98-99%, respectively. The reviews also showed that there was high heterogeneity in sensitivity which might be partly due to using varying protocols for stool processing, with differences in reagents and methods of homogenization and filtering (5). Moreover, most of the stool processing methods for Xpert testing were rather quite complex and cannot be performed at the lower levels of the healthcare system. This shows that there is a lack of standardized stool preparation and testing protocols and warrants the optimization and standardization of the stool processing methods that can be used at the decentralized level. The KNCV Tuberculosis Foundation (KNCV) and Ethiopian Public Health Institute (EPHI) developed the Simple One-Step (SOS) stool processing method, which can be applied in any GeneXpert laboratory (8).

This SOS stool processing method uses similar steps as sputum Xpert testing and does not require additional materials or equipment other than an applicator to pick the correct stool amount for testing. Since the method is as simple as sputum Xpert processing, it can be performed at all sites where a GeneXpert instrument is functional after providing minimal training to the staff involved in Xpert testing(8).

The SOS method has shown to be successful, with a low rate of unsuccessful results reported, in a demonstration study conducted in Ethiopia, in which multiple laboratories were involved (8). Furthermore, a head-to-head comparison study, in which the SOS method is compared to other stool processing methods showed similar sensitivity and specificity. The SOS stool method was most suitable for low-resource settings, because of its low error rate, processing time, and minimal requirements regarding biosafety precautions and laboratory equipment (9)

To gain more knowledge and in-depth experience on how the SOS stool processing method with Xpert-Ultra would behave if included in the routine diagnostic pathway for (childhood) TB and rolled out under the national TB program, we aim to further test and optimize the SOS processing method for the detection of TB in stool by Xpert-Ultra and its ability to tolerate perturbations (robustness). The study will also help to fine-tune standard operating procedures (SOPs) for the SOS method.

## Materials and Methods

### Design plan

#### Study setup and period

This will be a cross-sectional study that will consist of a series of experiments on consecutive stool samples collected from children and adults that are either sputum/NGA and/or stool Xpert-Ultra MTB positive. The study will be conducted in multiple health care facilities (>20) in Addis Ababa, Ethiopia. The facilities have a relatively high number of TB patients and have experience with participation in research. Children are being recruited for another related study, that assesses the diagnostic accuracy of Xpert stool testing using the SOS stool processing method, called Alternatives to Sputum Testing for Tuberculosis in Indonesia and Ethiopia (ASTTIE), see “S1 Figure”. Therefore, MTB positive children who will be identified in the ASTTIE study will also be used in the current study (PODTEC study). We will also recruit adults with MTB detected in sputum from the same facilities. The study was originally planned to be carried out till the end of 2020. However, due to the COVID-19 pandemic, the study period has been extended.

#### Study population

Children aged ≤10 years from the ASTTIE study who are sputum/NGA and/or stool Xpert-Ultra MTB positive, and consecutive sputum Xpert MTB-positive adults presenting in the selected health facilities will constitute the study population. Eligible persons, or their caregivers will be requested to sign (parental) consent or assent, depending on the age of the participant. The exclusion criteria include being critically sick i.e., those who are in coma, terminally ill due to chronic debilitating co-morbidities, or other conditions determined to be “critical” by the treating physician, being on TB treatment for longer than 5 days at the time of recruitment, and refusal to sign the informed consent.

#### Participant enrolment and stool collection

For children, the facility coordinator from the ASTTIE project will daily retrieve stool Xpert-Ultra results from the study sites and checks for eligibility. Parents of eligible children and eligible adults will be asked for informed consent for participation in this study, see “S1 appendix”. For children, this is an additional consent to the consent already provided for the ASTTIE study. After enrolment in the study, for children, the remainder of the initial stool sample (stool 1) will be collected and transported to EPHI. Participants will be provided with two (children) or three (adults) large stool containers to allow collection of at least 30 grams of stool. They will be instructed on how to collect and store the samples till delivery at the site.

Three appointments will be made to submit the additional stool samples. When the samples are submitted, information will be collected on the stool submission form “S2 appendix” about the date and time of collection at the household, storage conditions at the household and during transport and date and time of arrival at the site. To maximize the likelihood of finding MTB in the stool, the additional stool samples should be collected within 5 days after the participant’s TB treatment starts. Participants will be reimbursed for travel costs.

The stool samples will be kept in a cold chain at the site until they are transported to EPHI on the same day. The site will inform the study coordinator which will assign a dedicated transporter for this research purpose. The time between contacting the study coordinator and the pick-up of the samples is expected to be within 2 hours after delivery on-site, which means that the samples should arrive on the same day of collection at EPHI. After arrival at EPHI, the dedicated laboratory technician will be ready to receive and register the samples and start the cascade of experiments as shown in the sample flow diagram “S1 Figure 1”.

#### Design of the experiments

In total three experiments are designed and the lay out is depicted in “S2 Figure” and “S3 Figure”.

### Experiment 1: Stool sample strategy

This experiment will investigate if, and by how much, the positivity rate of Xpert-Ultra on stool increases when more than one sample from the same participant is tested. It also indicates how homogeneous the Mycobacteria are distributed within the stool samples and across different stools. Furthermore, it will provide insight in repeat testing if required due to unsuccessful test result, whether to advise to perform the repeat test from the same stool or from a fresh stool sample. This will be done by testing three stools from the same participant collected during consecutive days (Experiment 1a) as well as three aliquots (North, South and East/West) from the same stool sample (Experiment 1b). For children, two aliquots will be collected from the first stool sample “S1 Figure 1”. Aliquots will be tested using Xpert-Ultra, totaling a maximum of nine Xpert-Ultra tests per participant.

### Experiment 2: Stool sample storage conditions

This experiment will investigate how long and under which conditions stool can be kept without losing sensitivity to detect TB on Xpert-Ultra or increasing rates of unsuccessful tests. This is done by testing multiple aliquots taken from a known Xpert-Ultra MTB-positive stool sample after storing aliquots from that Xpert-Ultra MTB-positive stool at three temperatures; a) refrigerator 2-8°C, b) room temperature 20-22°C, and c) incubator 37°C and at four time periods; 2, 3, 5 and 10 days. These experiments will be done using aliquots remaining from the second and third stool samples collected for Experiment 1. These stool samples are expected to have more stools collected and probably the shortest storage time between collection at the site and preparation at EPHI.

### Experiment 3: Optimization and evaluating robustness of the SOS stool processing method

This experiment consists of a series of sub-experiments that will investigate if the SOS stool processing method can be further optimized to increase its recovery rate to detect TB. Although the SOS stool processing method is simple and contains minimal processing steps, certain steps might still be adapted to see if this influences the test’s sensitivity. This is done by testing multiple aliquots taken from a known Xpert-Ultra MTB-positive stool sample processed using slightly different approaches. The first sub-experiment (3a) varies the incubation time and shaking method during the processing of stool. The second sub-experiment (3b) assesses the optimum and maximum time and temperature for keeping the processed stool-sample reagent mixture before Xpert testing is conducted on the different incubation steps. The third sub-experiment (3c) assesses the optimum and maximum stool volume.

Stool consistency and bacterial load are two important factors that might influence the outcome of the experiments. Therefore, samples with different consistency and bacterial load will be included in all experiments.

If the Xpert-Ultra result is unsuccessful (i.e., the result is “invalid” or “error”), the test will NOT be repeated as this is part of the study outcome.

The SOS stool method’s comprehensive instructions can be found on KNCV website (10). In “S3 appendix” a schematic overview of the SOS stool method is provided. Depending on the stool consistency the protocol for solid stool or liquid stool is followed.

### Sampling plan

#### Variables and outcome measures

The primary outcome measure will be the Xpert-Ultra MTB quantitative result and positivity rate of stool specimens processed using the SOS stool processing method under the different experimental conditions will be compared to the baseline (on-site) Xpert-Ultra MTB positive test result of stool processed using the SOS stool processing method.

The secondary outcome measure will be the rate of Xpert-Ultra unsuccessful test results.

#### Sample size

At the selected health facilities, a maximum of 750 children with presumptive TB will be enrolled in ASTTIE during the recruitment period of this study. With Xpert-Ultra MTB-positivity rate of 5%, up to 32 children will be available for the optimization exercises. We aim to supplement this with up to 50 participants by also recruiting adults from the health facilities participating in ASTTIE, as described above. Thus, for the experiments, we will have stool samples for around 50 individuals available, totaling a maximum of 150 stool samples. “S4 Figure” indicates the minimum rates of conversion (Xpert MTB-to Xpert MTB+), respectively reversion (Xpert MTB+ to Xpert MTB-), that can be picked up with statistical significance with this sample size.

#### Analysis plan and data collection

Data will be collected on age and sex of the participant, TB suggestive symptoms and TB contact history, Xpert-Ultra result for the initial stool (children only) and sputum/NGA sample, date, and place (participant’s home or health facility) of stool of collection, date of stool receipt at the NTRL, stool storage and transport temperature until receipt at the NTRL, and stool consistency. Detailed information on the experiments’ conditions will also be collected as well as the cycle threshold (Ct) values for all probes and error codes in case of errors.

#### Data entry, storage, and management

Each stool sample will be submitted to EPHI together with a stool submission form “S2 appendix”. Details when conducting the experiment are collected on the experiment form “S4 appendix”. The forms are stored at EPHI. All data will be entered into pre-structured EpiData files (EpiData version 3.1; www.epidata.dk). A random 10% of the data will be re-entered in a separate file to check the quality of data entry. If more than 3% of errors are found in key variables (experiment conditions and Xpert result), full double data entry will be conducted.

#### Data interpretation

The main study outcome is the semi-quantitative Xpert Ultra result as provided by the GeneXpert instrument (trace, very low, low, medium, high, error, invalid or no result), interpreted as per the manufacturer’s instructions. The individual probes’ Ct values are the main quantitative study outcomes. We consider higher Ct values to represent lower bacillary loads, as these indicate that more PCR cycles are needed to reach the threshold of MTB detected. Error codes will be interpreted following the manufacturer’s guidance to understand the likely cause of the error.

#### Statistical analysis

Statistical analysis will be performed by the STATA/SE (version 15; StataCorp) statistical software package. The Xpert-Ultra stool result from each aliquot will be compared with the baseline Xpert-Ultra. Trends in the proportion of samples being MTB-positive and the proportion of samples with unsuccessful results over e.g., increasing storage time or temperature, or increasing amounts of stool added, will be analyzed using Wilcoxon-like test for trend of across ordered groups using nptrend (11). Logistic regression will be applied to assess factors associated with stool MTB positivity and unsuccessful test outcomes. We will assess if there is indication that stool samples are nested into individual participants, just as aliquots of one stool sample may be nested into individual stool samples, by comparing the outcomes of simple (multivariate) logistic regression to the outcomes of multilevel mixed-effects logistic regression using the likelihood-ratio test to determine the best model fit.

## Ethical considerations

The study obtained ethical clearance from the Review Boards of the Ethiopian Public Health Institute (EPHI-IRB) (Protocol no EPHI-IRB-234-2020). The project will follow the routine procedure of patients’ recruitment into studies, follow-up, and analysis as well as drawing of conclusions. Informed parental consent will be obtained from the children’s legal guardians. Participants’ information will be kept confidential, and the digital files used for analysis will only have the PODTEC laboratory code and ASTTIE unique person identification code (UPIC) and will not contain any names or other personal identifying information of the participant. Participants’ information will be kept confidential, and the digital files used for analysis will only have the PODTEC laboratory code and the ASTTIE UPIC and will not contain any names or other personal identifying information of the participant.

## Discussion

This is the first study protocol in which the sampling strategy and robustness of a stool processing method will be investigated. Based on the experiment’s findings, certain steps in the current SOP of the SOS stool method might be adjusted. The experiments will be performed using samples from presumptive TB cases, so for the patients for whom the test will be used in practice in a country with a relatively high TB burden. Uniquely, stool samples will not be spiked with mycobacteria as in other studies (9), (12). We expect that the MTB distribution is different in the stool samples from TB patients than in the spiked stools. Moreover, we will include children who have mostly have paucibacillary TB and who will benefit most from stool Xpert testing, as they cannot easily provide sputum. The study population will be drawn from locations where the test is expected to be conducted in the future, providing more realistic insights in the possibility of implementing the method in routine and collection of multiple samples.

The experiments are based on the controlled simulation of plausible scenarios, i.e. situations that can occur in practice, such as long transit times at high temperatures, long contact time of stool with the sample reagent before Xpert testing, variation in the stool portion size picked for processing, and variation in sample processing. The outcome will provide insights of the robustness of the method and will show how far certain steps can be stretched. It will provide practical outcomes that will enable the laboratory personnel and healthcare professionals involved in the stool testing to implement the most optimal protocol.

The main results will be presented both at local and international scientific meetings. The results will also be disseminated in the form of peer reviewed publications and as policy briefs. Key audiences for the dissemination will include global scientific advisory group members, local technical advisory committee (TAC) members and NTP. Study host communities will also be informed about the key results of the study through appropriate popular media.

## Data Availability

No datasets were generated or analysed during the current study. All relevant data from this study will be made available upon study completion.

## Acknowledgements

We would like to acknowledge the Ethiopian Public Health Institute and KNCV Tuberculosis Foundation. We would also like to thank all healthcare facilities that will participate in the study by recruiting the study participants. We would further like to acknowledge Mamush Sahile from KNCV Ethiopia for his assistance in the study.

## Author contribution

**Conceptualization: PdH, BY, AS, EWT, DJ**

**Data curation: BY, GD, MA, BS**

**Formal analysis: EWT, BY, PdH**

**Funding acquisition: EWT, PdH, DJ, AB**

**Investigation: PdH, BY, GD, AS, BS, EM, EWT, DJ**

**Methodology: PdH, BY, GD, MA, AS, BS, EM, ZGD, AK, MG, EWT, DJ**

**Project administration: DJ, BY, YD, BS, EM, ZGD, AB**

**Resources: DJ**

**Software: EWT**

**Supervision: PdH, EWT, DJ, BY, GD, MA, AS, YD, BS, EM, ZGD, AK, MG, AB**

**Validation: EWT**

**Visualization: EWT, PdH**

**Writing-original draft: DJ, PdH, EWT, BY**

**Writing-review & editing: all authors**

---

**Figure 1:**
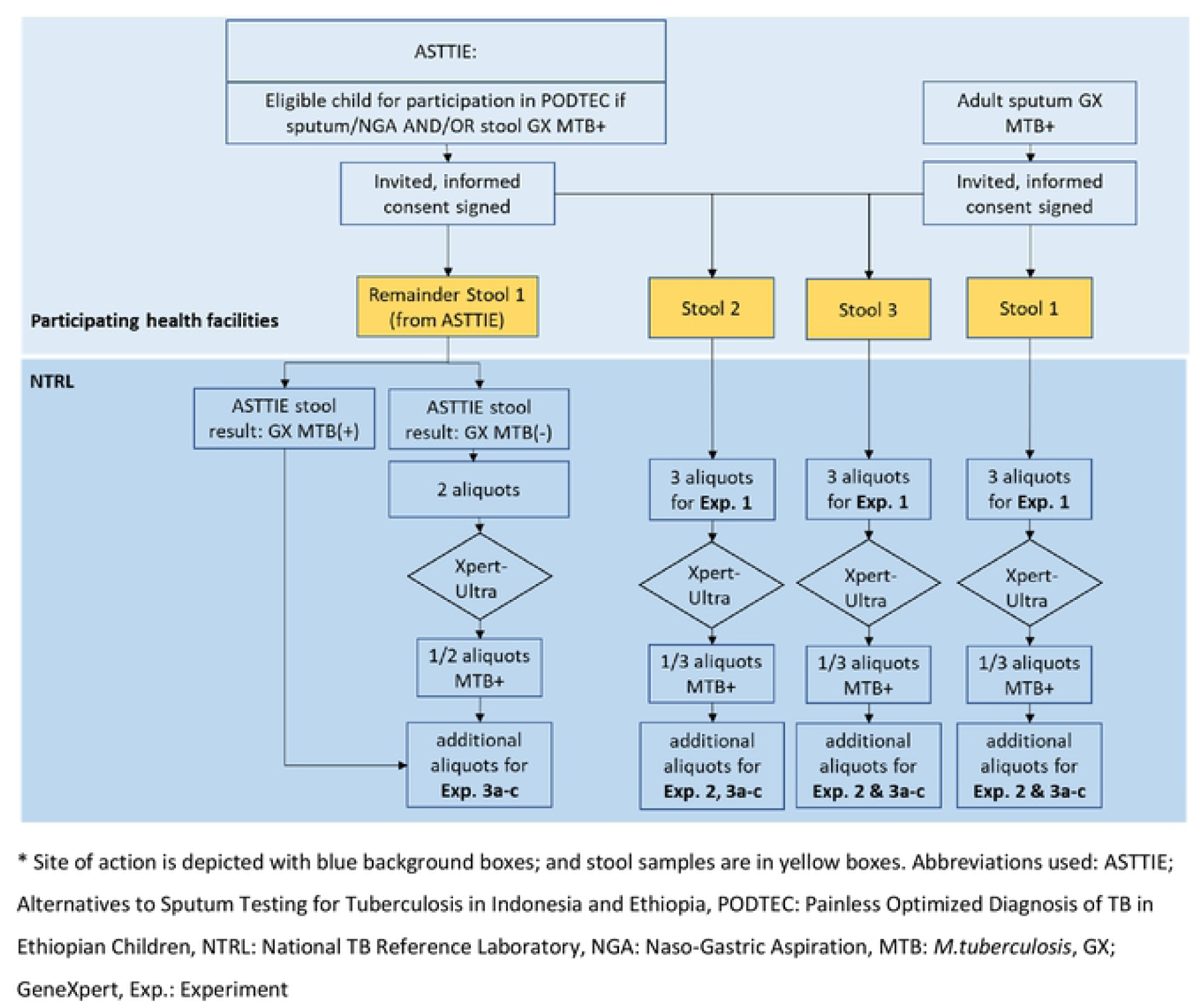
Study flow diagram representing steps from inclusion of adults and children to arrival of the stool samples at EPHI and their allocation to the different experiments

**Figure 2.**
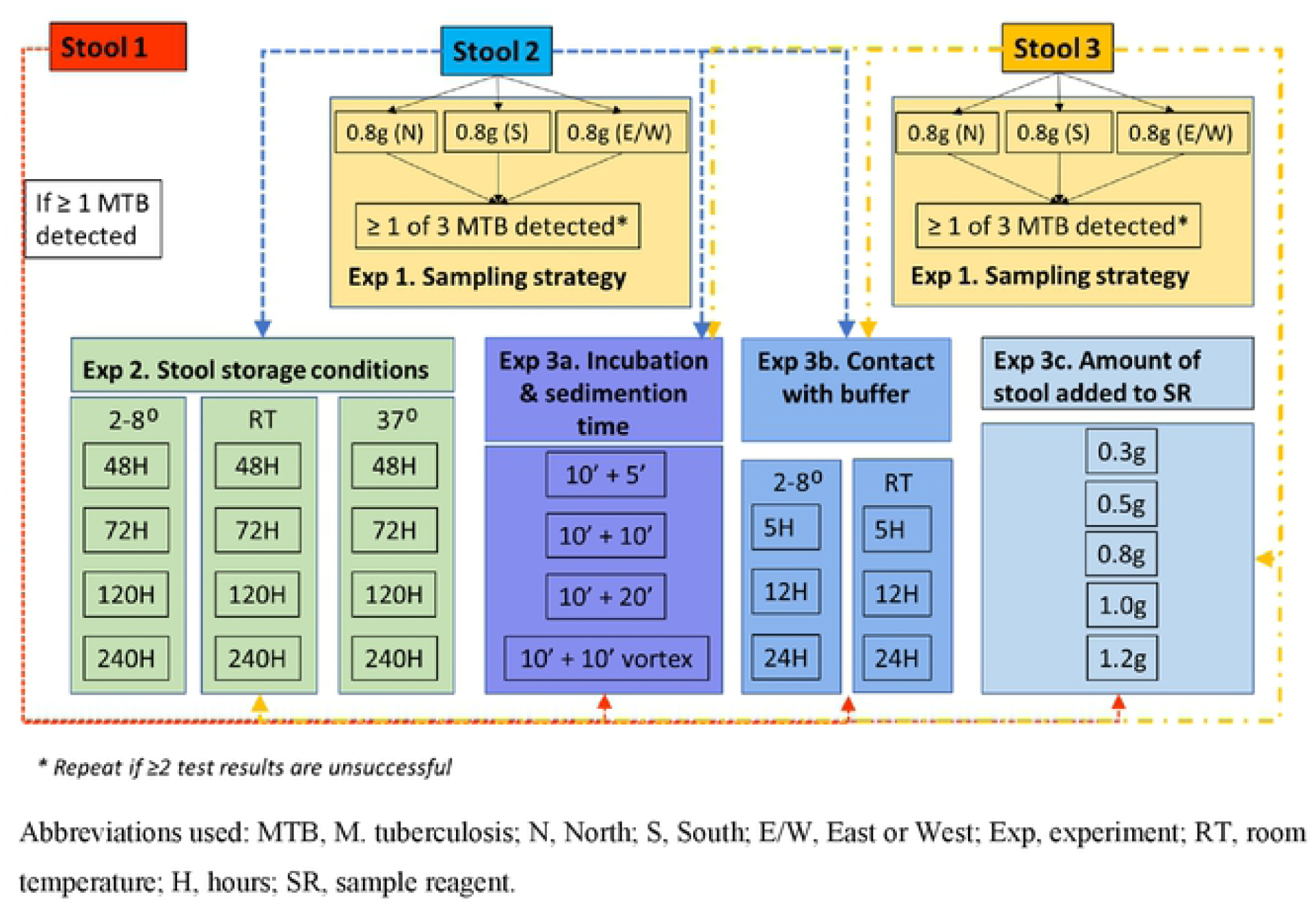
Overview, of experiments. Arrows indicate what experiments are done, vith the different stool specirnens. Note that for adult participants, in principle all experiments will be done on all three stool samples, provided that enough stool is collected per bowel movement.

**Figure 3.**
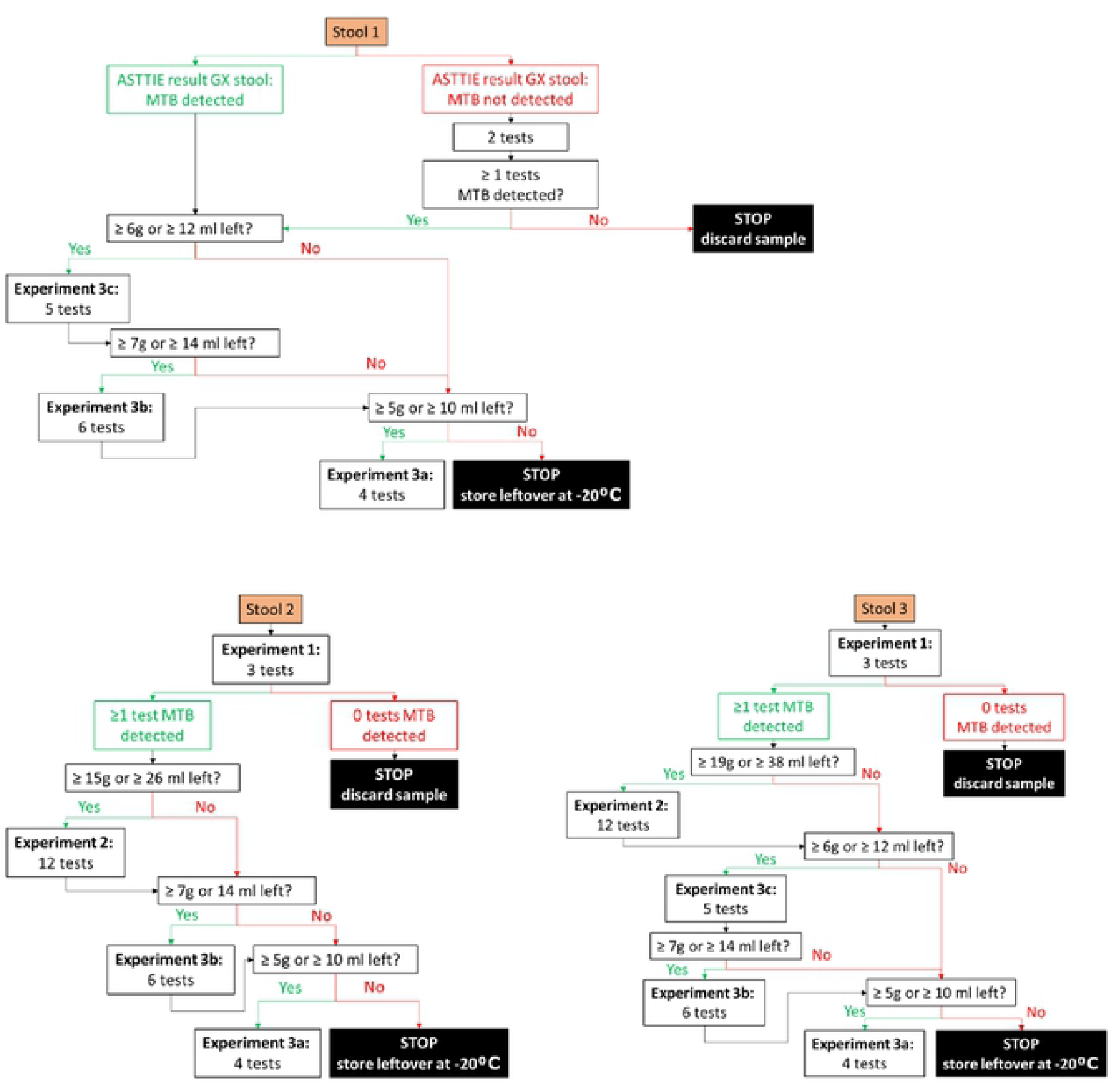
Overview of assigning Stool to the Experiments outlined in figure 2. Note that for adults, in principle, all experiments will be conducted if there is enough stool available per sample.

**Figure 4:**
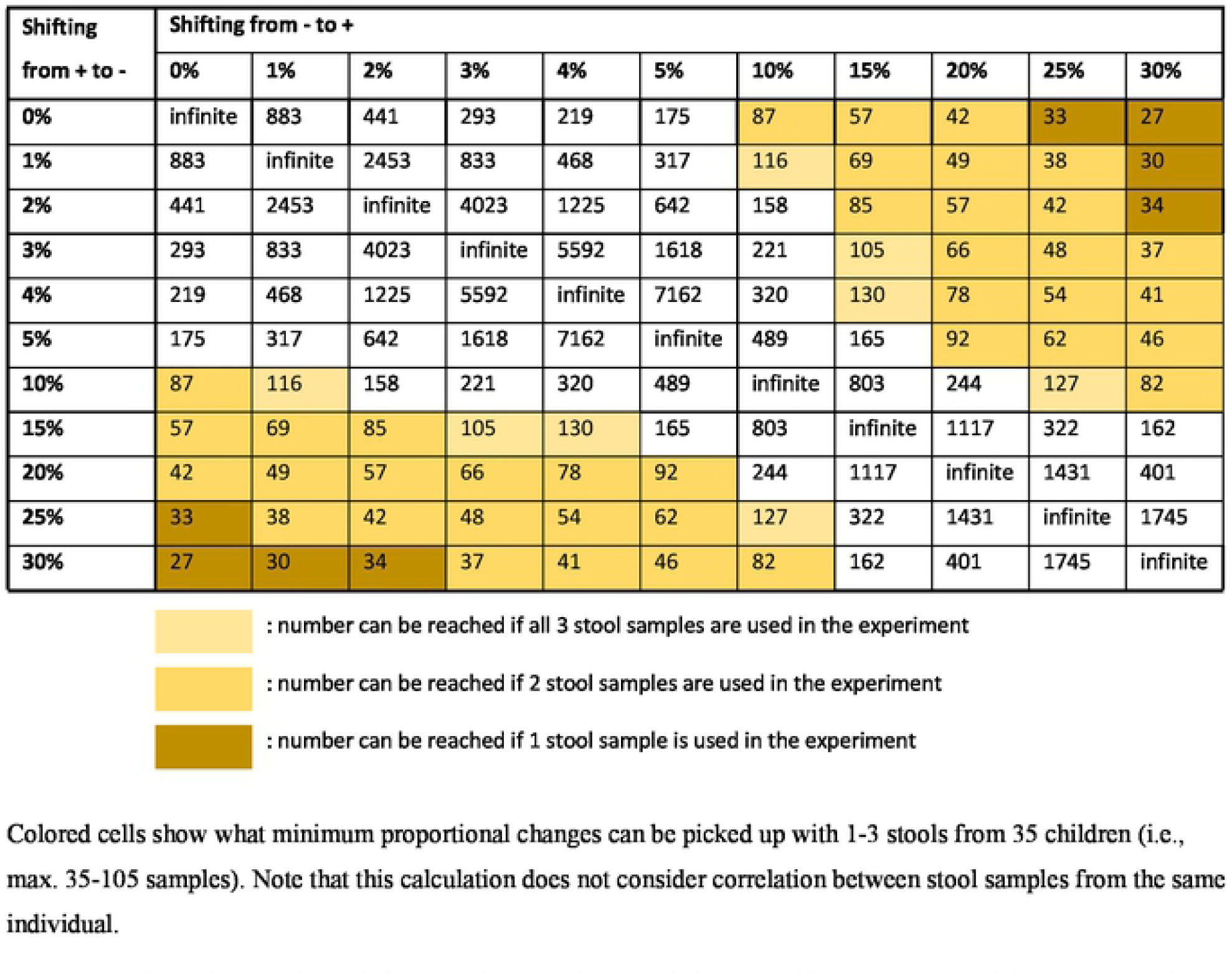
Sample size by minimum changes in positivity rate (from MTB-positive to -negative, and from MTB-negative to -positive Xpert-Ultra result), calculated using the Statulator; (11).

